# Socioeconomic Determinants of Guideline-Concordant Therapy for Early-Stage Non-Small Cell Lung Cancer: A Population-Based Analysis from Appalachian and Non-Appalachian Ohio, 2004-2015

**DOI:** 10.64898/2026.06.20.26356121

**Authors:** Jeremiah T. Martin, Wendi Waugh

**Author notes:** Corresponding author. (Jeremiah T. Martin). Prior presentation: An earlier version of this work was presented in abstract form at the Society of Thoracic Surgeons 56th Annual Meeting, New Orleans, Louisiana, January 28, 2020.

## Abstract

**Purpose:** To examine the relative contributions of insurance, county-level poverty, and other socioeconomic factors, as compared with Appalachian geography, to receipt of guideline-concordant therapy for early-stage non-small cell lung cancer (NSCLC) in Appalachian and non-Appalachian Ohio.

**Methods:** Retrospective population-based cohort study using the Ohio Cancer Incidence Surveillance System. We identified adults diagnosed with early-stage NSCLC between 2004 and 2015 (N=26,756). The primary outcome was receipt of guideline-concordant local therapy (surgery or definitive radiation). Rural-urban classification used USDA Rural-Urban Continuum Codes. Multivariable logistic regression and Cox proportional hazards models assessed predictors of treatment and survival, with E-values, race-stratified models, and propensity score weighting as sensitivity analyses.

**Findings:** Median age was 71 years; 50.3% were male, 83.8% non-Hispanic White, and 20.4% Appalachian. Overall, 83.6% received guideline-concordant local therapy (59.6% surgery, 24.0% radiation). In adjusted analysis, Medicaid (adjusted odds ratio [OR] 0.53, 95% confidence interval [CI] 0.44-0.63; adjusted risk ratio [RR] 0.94, 0.91-0.96), county-level poverty >20% (OR 0.77, 95% CI 0.68-0.87; RR 0.96, 0.95-0.98), and unmarried status were independently associated with lower therapy receipt, whereas Appalachian residence was associated with modestly higher receipt (OR 1.17, 95% CI 1.06-1.29; RR 1.02, 1.01-1.04). Therapy rates converged across regions over the study period (year × Appalachian interaction p<0.001). Mortality was independently associated with lack of local therapy (adjusted hazard ratio [HR] 4.33, 95% CI 4.10-4.56), Medicaid (HR 1.25, 95% CI 1.14-1.37), and poverty >20% (HR 1.13, 95% CI 1.07-1.20).

**Conclusions:** Socioeconomic factors, particularly Medicaid insurance and county-level poverty, were the patient characteristics most strongly associated with lower receipt of guideline-concordant therapy, whereas Appalachian residence was not a barrier. Findings support targeted interventions addressing insurance-related and poverty-related barriers to lung cancer care in high-poverty communities regardless of geographic designation.

## INTRODUCTION

Lung cancer remains the leading cause of cancer-related mortality in the United States, with an estimated 234,580 new cases and 125,070 deaths in 2024.[1] For patients with early-stage non-small cell lung cancer (NSCLC), surgical resection is the standard of care, achieving 5-year survival rates of 60-80% for stage I disease.[2] Stereotactic body radiation therapy (SBRT) has emerged as a guideline-concordant alternative for medically inoperable or selected operable patients, with randomized trials demonstrating durable local control and survival outcomes comparable to surgery in carefully selected populations.[3,4] Despite strong evidence supporting definitive local therapy, a substantial proportion of patients with potentially curable disease fail to receive guideline-concordant treatment. A landmark National Cancer Database (NCDB) analysis of 441,812 patients found that only 62% received NCCN-recommended therapy, with markedly lower rates among elderly and non-Hispanic Black patients.[5]

Socioeconomic and geographic disparities in lung cancer treatment have been consistently documented. Stokes et al. demonstrated that patients insured by Medicaid (OR 0.53) or uninsured (OR 0.50) were significantly less likely to undergo surgical resection for early-stage NSCLC,[6] while Ebner et al. showed that patients with multiple socioeconomic risk factors had a 4.7-fold increase in the odds of receiving no cancer-directed therapy.[7] Rural residence further compounds these barriers: Logan et al. reported that rural patients had 21% higher adjusted odds of not receiving surgery, driven by both lower rates of surgical recommendation and lower receipt even when surgery was recommended.[8] However, national datasets commonly used to examine these disparities have important limitations. SEER captures only 34.6% of the U.S. population and does not include Ohio, while the NCDB includes only patients treated at Commission on Cancer-accredited facilities, potentially excluding individuals who never access specialized cancer care.[9]

The Appalachian region spans 423 counties across 13 states and is characterized by persistent poverty, limited healthcare access, higher smoking prevalence, and excess cancer mortality.[10–12] Appalachian Kentucky has been identified as a lung cancer “hotspot,” with incidence rates up to 3.5 times the national average and worse stage-specific survival.[13] In Tennessee, residents of Appalachian counties had 35% lower odds of receiving surgical treatment for localized lung cancer compared with those in non-Appalachian counties.[14] Within Ohio, a study of the state’s poorest counties, nearly all designated Appalachian, demonstrated 19% higher all-cancer mortality, higher rates of late-stage diagnosis, and greater rates of no treatment compared with more affluent counties.[15] Nevertheless, it remains uncertain whether these observed disparities are attributable to Appalachian geography itself or instead reflect the concentration of socioeconomic disadvantage within Appalachian counties.

Ohio provides a distinct opportunity to disentangle these effects. Of its 88 counties, 32 are designated Appalachian by the Appalachian Regional Commission, enabling within-state comparisons that inherently control for variation in state-level policies related to Medicaid expansion, tobacco control, and cancer screening.[16] The Ohio Cancer Incidence Surveillance System (OCISS), a comprehensive population-based registry capturing all cancer diagnoses regardless of treatment location, offers near-complete coverage that overcomes key limitations of national databases. In addition, the availability of the NAACCR “Reason No Surgery” variable, rarely examined in population-based analyses,[17,18] allows for mechanistic assessment of why eligible patients do not undergo surgical resection.

To our knowledge, no prior study has used a single-state population-based cancer registry to formally decompose the contributions of insurance, county-level poverty, race, and rurality to receipt of guideline-concordant local therapy in early-stage NSCLC with explicit testing of geographic and temporal interactions across an Appalachian/non-Appalachian divide. Accordingly, we sought to determine whether Appalachian residence is independently associated with rates of guideline-concordant local therapy after adjustment for socioeconomic factors, and to quantify the relative contributions of insurance status, county-level poverty, race, rurality, and other factors to treatment patterns and survival outcomes across Appalachian and non-Appalachian regions of Ohio. We hypothesized a priori that socioeconomic factors, particularly Medicaid insurance and high county-level poverty, would be independently associated with lower receipt of therapy, that Appalachian residence would show little or no independent association after adjustment for these factors, and that any geographic difference would converge over time concurrent with Ohio’s 2014 Medicaid expansion and the national diffusion of stereotactic body radiation therapy for early-stage disease.

## METHODS

### Study Design and Data Source

We conducted a retrospective, population-based cohort study using the Ohio Cancer Incidence Surveillance System (OCISS), the statewide cancer registry that captures all cancer diagnoses among Ohio residents regardless of where treatment is received, in accordance with Ohio Revised Code §3701.262.[19] OCISS adheres to North American Association of Central Cancer Registries (NAACCR) coding standards and achieves greater than 95% case completeness.

This study was reviewed and approved under expedited review by the Ohio Department of Health Human Subjects Institutional Review Board (FWA00001963, IRB00002180; Protocol 2018-01), with continuing review through February 14, 2020. The de-identified analytic dataset was obtained from the Ohio Cancer Incidence Surveillance System under a signed Data Use Agreement and was first accessed for research in 2018; the authors did not have access to information that could identify individual participants. Because the analysis used de-identified secondary registry data, informed consent was not obtained from individual participants. We report this study in accordance with the Strengthening the Reporting of Observational Studies in Epidemiology (STROBE) statement; the completed checklist is provided as S1 Checklist.

### Study Population

We identified all patients aged ≥18 years diagnosed with lung cancer (ICD-O-3 site codes C34.0-C34.9) between January 1, 2004, and December 31, 2015. We restricted the cohort to non-small cell lung cancer (NSCLC) by excluding small cell histologies (ICD-O-3 morphology codes 8041-8045). Early-stage disease was defined using the SEER Summary Stage variable (OHStage), including in situ (0), localized (1), and regional disease with direct extension only (2).

We excluded cases ascertained solely by autopsy or death certificate, tumors with non-malignant behavior codes, and patients with zero or missing survival time. The start date of 2004 was selected to coincide with the availability of Collaborative Staging variables, and follow-up was truncated at 2015 to ensure a minimum of two years of potential observation for survival analyses.

The final analytic cohort comprised 26,756 patients (Fig 1).

**Fig 1.**
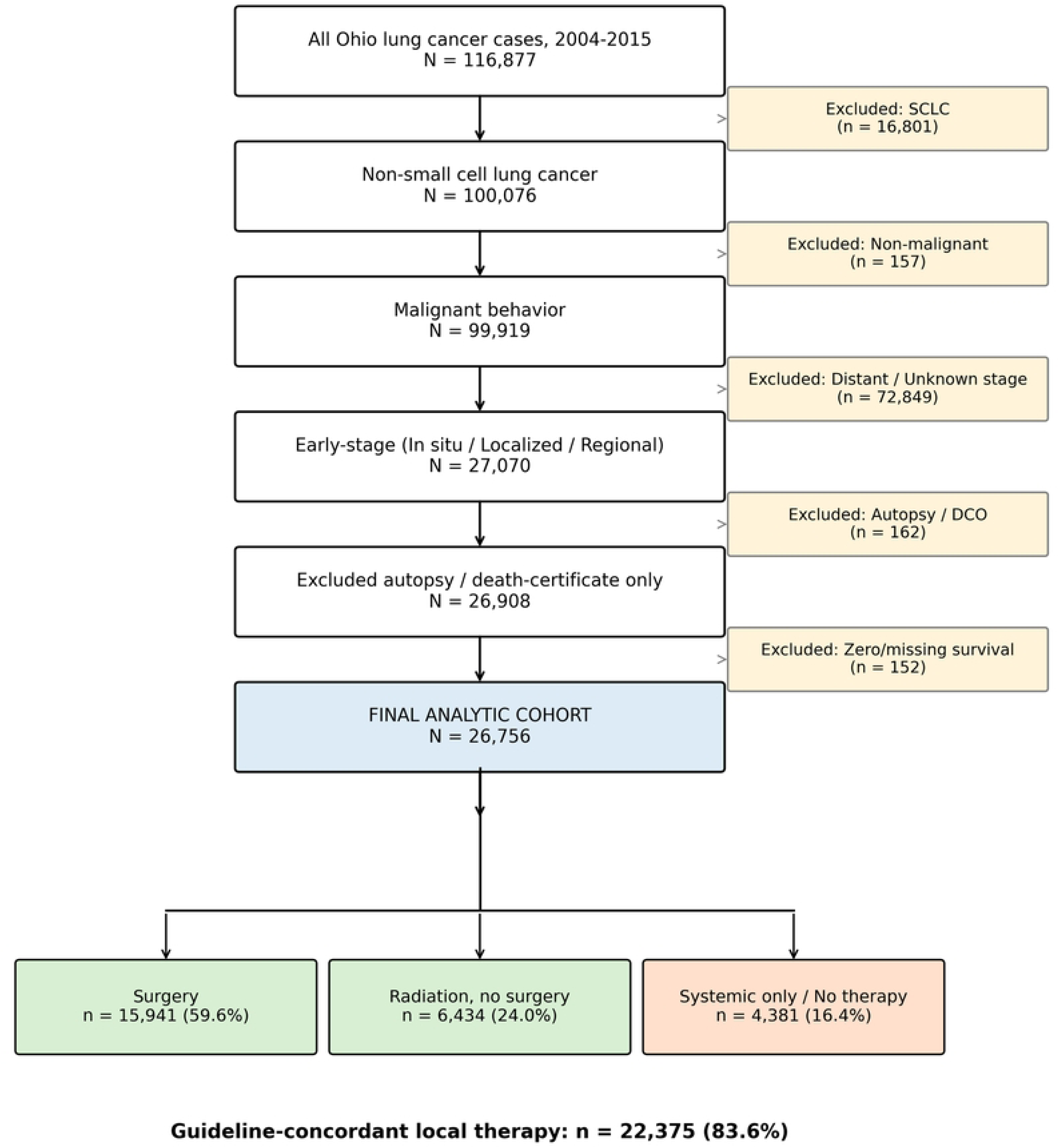
Cohort Selection Flow Diagram - Patient Selection from OCISS, 2004-2015.

### Variable Definitions

#### Primary outcome

Guideline-concordant local therapy was defined as receipt of surgery (NAACCR RxSumSurgPSite codes 20-98) or definitive radiation therapy (NAACCR RxSumRad codes 1-8). This binary variable served as the primary outcome for logistic regression models.

#### Treatment categorization

Patients were classified into four mutually exclusive treatment categories: (1) Surgery (codes 20-98); (2) Radiation without surgery; (3) Systemic therapy only (chemotherapy, immunotherapy, or hormonal therapy without local therapy); (4) No cancer-directed therapy. **Primary exposure:** Appalachian status was defined using the Appalachian Regional Commission’s designation of 32 Ohio counties as Appalachian.[16] Patients were classified as Appalachian or non-Appalachian based on county of diagnosis.

#### Covariates

Age at diagnosis (continuous); sex; race/ethnicity (non-Hispanic White, non-Hispanic Black, Hispanic, Other/Unknown); marital status (married/partnered, single, divorced/separated, widowed); insurance at diagnosis (private, Medicare, Medicaid, uninsured/self-pay, Military/VA, unknown); county-level poverty index (<5%, 5-10%, 10-20%, >20%; NAACCR PovertyInd); rural-urban classification using USDA Rural-Urban Continuum Codes (RUCC),[20] categorized as Metro (RUCC 1-3), Adjacent to metro (RUCC 4, 6, 8), and Rural (RUCC 5, 7, 9); SEER Summary Stage (localized, regional with direct extension); tumor size (continuous, mm); tumor grade (well, moderately, poorly differentiated, undifferentiated, unknown); and smoking history (never, former, current, unknown).

#### Reason for no surgery

Among patients not undergoing resection, the NAACCR ReasonNoSurg variable was categorized as: surgery not recommended, contraindicated due to comorbidity, patient refused, died before treatment, recommended but unknown if performed, and unknown/not applicable.

#### Survival

Overall survival was calculated from diagnosis date to date of last contact or death. Vital status was ascertained from death certificates and active follow-up. Cancer-specific survival used the underlying cause of death variable (DthCause). Survival was capped at 60 months.

### Statistical Analysis

Descriptive statistics were calculated for all study variables, overall and stratified by Appalachian status (Table 1) and by treatment category. Continuous variables were summarized using medians with interquartile ranges (IQRs), and categorical variables were reported as frequencies with percentages. Between-group comparisons were conducted using the Wilcoxon rank-sum test for continuous variables and chi-squared tests for categorical variables.

**Table 1.** Patient and Tumor Characteristics by Appalachian Status (N=26,756)

| Variable | Level | Overall<br>(N=26,756) | Non-Appalachian<br>(N=21,298) | Appalachian<br>(N=5,458) | p |
| --- | --- | --- | --- | --- | --- |
| Age, years | Median (IQR) | 71 (63-77) | 71 (63-78) | 70 (63-76) | <0.001 |
| Sex | Male | 13,464 (50.3%) | 10,501 (49.3%) | 2,963 (54.3%) | <0.001 |
| Female | 13,291 (49.7%) | 10,796 (50.7%) | 2,495 (45.7%) |  |  |
| Race/Ethnicity | NH White | 22,422 (83.8%) | 17,592 (82.6%) | 4,830 (88.5%) | <0.001 |
| NH Black | 2,571 (9.6%) | 2,390 (11.2%) | 181 (3.3%) |  |  |
| Hispanic | 91 (0.3%) | 82 (0.4%) | 9 (0.2%) |  |  |
| Other/Unknown | 1,672 (6.2%) | 1,234 (5.8%) | 438 (8%) |  |  |
| Marital Status | Married/Partnered | 13,831 (53.2%) | 10,905 (52.6%) | 2,926 (55.3%) | <0.001 |
| Single | 2,774 (10.7%) | 2,320 (11.2%) | 454 (8.6%) |  |  |
| Divorced/Separated | 3,644 (14%) | 2,894 (14%) | 750 (14.2%) |  |  |
| Widowed | 5,766 (22.2%) | 4,603 (22.2%) | 1,163 (22%) |  |  |
| Insurance | Private | 4,789 (18.2%) | 3,995 (19.1%) | 794 (14.8%) | <0.001 |
| Medicare | 17,673 (67.3%) | 14,009 (67.1%) | 3,664 (68.3%) |  |  |
| Medicaid | 1,571 (6%) | 1,229 (5.9%) | 342 (6.4%) |  |  |
| Uninsured/Self-pay | 1,211 (4.6%) | 884 (4.2%) | 327 (6.1%) |  |  |
| Military/VA | 400 (1.5%) | 293 (1.4%) | 107 (2%) |  |  |
| Unknown | 615 (2.3%) | 481 (2.3%) | 134 (2.5%) |  |  |
| Poverty Index | <5% | 4,361 (16.3%) | 4,052 (19.1%) | 309 (5.7%) | <0.001 |
| 5-10% | 6,853 (25.7%) | 5,658 (26.6%) | 1,195 (22%) |  |  |
| 10-20% | 8,555 (32.1%) | 6,246 (29.4%) | 2,309 (42.4%) |  |  |
| >20% | 6,906 (25.9%) | 5,275 (24.8%) | 1,631 (30%) |  |  |
| Rural-Urban | Metro | 21,703 (81.1%) | 18,598 (87.3%) | 3,105 (56.9%) | <0.001 |
| Adjacent to metro | 4,564 (17.1%) | 2,492 (11.7%) | 2,072 (38%) |  |  |
| Rural | 489 (1.8%) | 208 (1%) | 281 (5.1%) |  |  |
| Stage | Localized | 19,240 (71.9%) | 15,397 (72.3%) | 3,843 (70.4%) | 0.006 |
| Regional (direct ext.) | 7,516 (28.1%) | 5,901 (27.7%) | 1,615 (29.6%) |  |  |
| Tumor Size, mm | Median (IQR) | 25 (17-39) | 25 (17-40) | 25 (17-39) | 0.636 |
| Treatment | Surgery | 15,941 (59.6%) | 12,614 (59.2%) | 3,327 (61%) | <0.001 |
| Radiation (no surgery) | 6,434 (24%) | 5,143 (24.1%) | 1,291 (23.7%) |  |  |
| Systemic only | 753 (2.8%) | 643 (3%) | 110 (2%) |  |  |
| No cancer-directed therapy | 3,628 (13.6%) | 2,898 (13.6%) | 730 (13.4%) |  |  |
| GC Local Therapy | Yes | 22,375 (83.6%) | 17,757 (83.4%) | 4,618 (84.6%) | 0.029 |
| No | 4,381 (16.4%) | 3,541 (16.6%) | 840 (15.4%) |  |  |

GC = guideline-concordant. Values are n (%) or median (IQR). Insurance percentages are computed over patients with non-missing payer information; 497 patients (1.9%) with missing insurance are not shown as a separate row.

#### Logistic regression

Multivariable logistic regression was used to evaluate predictors of receipt of guideline-concordant local therapy. Covariates were selected a priori using the Andersen behavioral model of healthcare utilization,[21] grouping variables into predisposing characteristics (age, sex, race/ethnicity, marital status), enabling resources (insurance, county-level poverty, rural-urban classification), and need (stage, tumor size, grade, smoking history), rather than through data-driven variable selection. Because the outcome prevalence was high (approximately 84%), the adjusted odds ratio overstates the corresponding risk ratio; logistic regression was retained as the primary model for direct comparability with the prior NSCLC treatment-disparities literature, which uniformly reports odds ratios,[6,17,18] and a modified-Poisson sensitivity analysis with robust standard errors[22] was performed to estimate adjusted risk ratios (S1 Table). To express adjusted associations on the absolute scale, we computed model-standardized predicted probabilities and risk differences (g-computation) for the principal socioeconomic exposures. Model performance and diagnostics were assessed using the C-statistic (area under the receiver operating characteristic curve), the Hosmer-Lemeshow goodness-of-fit test, and variance inflation factors (VIF) with a pre-specified concern threshold of VIF >5. Four interaction terms were tested a priori, each motivated by a specific hypothesis: poverty × race (effect modification of the SES gradient by race, given prior reports of differential SES gradients between Black and White cancer patients[23]); poverty × insurance status (whether financial barriers to care interact with payor source); Appalachian × poverty (whether the SES gradient differed by geographic region); and Appalachian × year of diagnosis (temporal convergence hypothesis, given Ohio’s 2014 Medicaid expansion[24] and the national diffusion of stereotactic body radiation therapy during the study period[25]). Interactions were tested using likelihood ratio tests at a significance threshold of α=0.10. Given this liberal threshold and the testing of four interaction terms, the temporal (Appalachian by year) interaction, which corresponds to the a priori convergence hypothesis, was treated as the primary confirmatory interaction; the remaining three were considered exploratory and hypothesis-generating.

A secondary multivariable logistic regression model examined predictors of receipt of surgery versus radiation among patients who received definitive local therapy.

#### Survival analysis

Kaplan-Meier curves with log-rank tests compared survival across treatment categories, Appalachian status, and poverty levels. Cox proportional hazards models estimated adjusted hazard ratios for all-cause mortality, including treatment category as a covariate because treatment is strongly associated with survival. The proportional hazards assumption was tested using Schoenfeld residuals; because the global test indicated departures from proportionality (Results), hazard ratios are interpreted as effects averaged over the 60-month follow-up. Cancer-specific mortality was additionally examined with competing-risks methods (cause-specific hazards and Fine-Gray subdistribution models), treating non-cancer death as a competing event. Additional Cox models included a 6-month landmark analysis (excluding early deaths to address immortal time bias),[26] cancer-specific survival (censoring non-cancer deaths), and a model restricted to patients receiving guideline-concordant therapy.

#### Missing data

Smoking history was missing for 59.3% and tumor grade for 28.9% of patients. Rather than excluding these patients (which would eliminate the majority of the cohort), we created an “Unknown” category for each variable and included it in models. Tumor size (6.6% missing) was imputed with the median. A sensitivity analysis using strict complete-case analysis (N=7,208) was performed to assess robustness. Insurance and marital status had <3% missing data each; the multivariable logistic regression model used complete cases for these covariates, excluding 1,304 patients (4.9% of the cohort) and yielding an analytic sample of N=25,452. To assess robustness to the Unknown-category approach for smoking and grade, we additionally fitted a multiple-imputation model (multivariate imputation by chained equations, m=10).

#### Sensitivity analyses

(1) E-values quantified the minimum strength an unmeasured confounder would need to explain away observed associations.[27,28] (2) Restriction to 2010-2015 tested robustness to contemporary treatment patterns. (3) Race-stratified models examined whether poverty effects differed between non-Hispanic White and non-Hispanic Black patients. (4) Exclusion of patients with documented medical contraindications to surgery (ReasonNoSurg codes indicating comorbidity or death before treatment) tested whether disparities persisted among potentially surgical candidates. (5) Inverse probability of treatment weighting (IPTW), using stabilized weights trimmed at the 1st and 99th percentiles, estimated the IPTW-adjusted association between guideline-concordant therapy and survival. (6) Temporal trend analysis with year × Appalachian interaction terms assessed whether disparities changed over time. All analyses were performed in R version 4.5.1 (R Foundation for Statistical Computing, Vienna, Austria). Two-sided p-values <0.05 were considered statistically significant.

### Use of Artificial Intelligence Tools

The authors conceived and directed this study, abstracted the data, and conducted the statistical analyses in R on a local computer. During preparation of the manuscript, the authors used a generative AI assistant (Claude Opus 4.8; Anthropic, PBC) to help write and refine analysis and figure code, to help draft and edit text, and to check the internal consistency of reported numbers. All analyses were executed locally and all outputs were verified by the authors against the source cancer-registry data. The authors reviewed and edited all AI-assisted code and text and take full responsibility for the integrity, accuracy, and validity of the work; the study’s results, interpretations, and conclusions are the authors’ own.

## RESULTS

### Study Population

Among 116,877 lung cancer cases diagnosed in Ohio between 2004 and 2015, we identified 26,756 patients with early-stage NSCLC meeting inclusion criteria (Fig 1). The median age at diagnosis was 71 years (IQR 63-77), 50.3% were male, and 83.8% were non-Hispanic White. Appalachian counties contributed 5,458 patients (20.4% of the cohort). Over a median follow-up of 40.3 months (reverse Kaplan-Meier method), 15,141 deaths (56.6%) occurred, of which 9,226 were attributed to cancer.

Baseline characteristics differed by Appalachian status (Table 1). Compared with non-Appalachian patients, those residing in Appalachian counties were slightly younger (median 70 vs 71 years, p<0.001), more often male (54.3% vs 49.3%), more frequently non-Hispanic White (88.5% vs 82.6%), and more likely to be current smokers (20.9% vs 17.7%). Appalachian patients were substantially more likely to live in high-poverty counties (>20% poverty: 30.0% vs 24.8%) and in non-metropolitan areas (43.1% vs 12.7%). Insurance coverage also differed, with Appalachian patients less likely to have private insurance (14.8% vs 19.1%) and more likely to be uninsured (6.1% vs 4.2%).

### Treatment Patterns

Overall, 83.6% of patients received guideline-concordant local therapy: 59.6% underwent surgical resection and 24.0% received radiation therapy without surgery. An additional 2.8% received systemic therapy alone, while 13.6% received no cancer-directed therapy. In unadjusted analyses, Appalachian patients had slightly higher rates of guideline-concordant therapy compared with non-Appalachian patients (84.6% vs 83.4%, p=0.029), as well as modestly higher surgical resection rates (61.0% vs 59.2%).

A clear socioeconomic gradient was observed: guideline-concordant therapy rates declined stepwise with increasing county-level poverty, from 85.3% in counties with <5% poverty to 83.5% in 5-10%, 82.8% in 10-20%, and 81.6% in >20% poverty counties (p<0.001).

### Reasons for No Surgery

Among 10,815 patients not undergoing resection, documented reasons were available for the majority. Surgery was not recommended in 73.6% of non-surgical patients, was contraindicated due to comorbidities in 14.3%, and was refused by 4.8%. Patient refusal rates were similar between Appalachian (5.1%) and non-Appalachian (4.7%) patients. Refusal rates differed modestly by race: 4.6% among non-Hispanic White patients and 5.7% among non-Hispanic Black patients.

### Predictors of Guideline-Concordant Therapy (Table 2)

In multivariable logistic regression (N=25,452, C-statistic=0.75), several factors were independently associated with lower odds of receiving guideline-concordant local therapy:

- **Insurance:** Medicaid (OR 0.53, 95% CI 0.44-0.63) and uninsured (OR 0.77, 95% CI 0.63-0.96) versus private insurance
- **Poverty:** County-level poverty >20% (OR 0.77, 95% CI 0.68-0.87), 10-20% (OR 0.87, 95% CI 0.78-0.98) versus <5%
- **Marital status:** Single (OR 0.57, 95% CI 0.51-0.65), widowed (OR 0.63, 95% CI 0.58-0.70), and divorced (OR 0.75, 95% CI 0.67-0.84) versus married
- **Age:** OR 0.95 per year (95% CI 0.95-0.96)
- **Tumor size:** Larger tumors associated with lower therapy receipt (OR 0.998 per mm) Female sex was associated with higher therapy receipt (OR 1.22, 95% CI 1.13-1.31). After adjustment for poverty, insurance, and other socioeconomic factors, Appalachian residence was associated with modestly higher therapy receipt (OR 1.17, 95% CI 1.06-1.29; p=0.002), consistent with its slightly higher unadjusted rate (84.6% vs 83.4%). Appalachian residence was therefore not a barrier to guideline-concordant therapy in this cohort; where lower receipt occurred, it was associated with the socioeconomic factors concentrated in these regions rather than with Appalachian geography itself.

**Table 2.** Adjusted Odds Ratios for Receipt of Guideline-Concordant Local Therapy (N=25,452)

| Group | Variable | OR | 95% CI | P |
| --- | --- | --- | --- | --- |
| Demographics | Age (per year) | 0.95 | 0.95-0.96 | <0.001 |
| Demographics | Female (ref: Male) | 1.22 | 1.13-1.31 | <0.001 |
| Race/Ethnicity | NH Black (ref: NH White) | 0.98 | 0.87-1.12 | 0.801 |
| Race/Ethnicity | Hispanic | 1.12 | 0.59-2.30 | 0.745 |
| Race/Ethnicity | Other/Unknown | 0.86 | 0.74-1.00 | 0.042 |
| Marital Status | Single (ref: Married) | 0.57 | 0.51-0.65 | <0.001 |
| Marital Status | Divorced/Separated | 0.75 | 0.67-0.84 | <0.001 |
| Marital Status | Widowed | 0.63 | 0.58-0.70 | <0.001 |
| Insurance | Medicare (ref: Private) | 0.88 | 0.78-0.99 | 0.043 |
| Insurance | Medicaid | 0.53 | 0.44-0.63 | <0.001 |
| Insurance | Uninsured/Self-pay | 0.77 | 0.63-0.96 | 0.018 |
| Insurance | Military/VA | 1.25 | 0.91-1.75 | 0.181 |
| Poverty Index | 5-10% (ref: <5%) | 0.89 | 0.80-1.00 | 0.061 |
| Poverty Index | 10-20% | 0.87 | 0.78-0.98 | 0.019 |
| Poverty Index | >20% | 0.77 | 0.68-0.87 | <0.001 |
| Geography | Appalachian (ref: Non-Appalachian) | 1.17 | 1.06-1.29 | 0.002 |
| Rural-Urban | Adjacent to metro (ref: Metro) | 0.86 | 0.78-0.95 | 0.003 |
| Rural-Urban | Rural | 0.87 | 0.67-1.15 | 0.330 |
| Stage | Regional (ref: Localized) | 0.91 | 0.84-0.98 | 0.018 |
| Tumor | Tumor size (per mm) | 1.00 | 1.00-1.00 | <0.001 |

*C-statistic = 0.75. OR = odds ratio; CI = confidence interval.*

Non-Hispanic Black race was not significantly associated with guideline-concordant therapy receipt after adjustment (OR 0.98, 95% CI 0.87-1.12, p=0.80).

Variance inflation factors were all <1.53, indicating no multicollinearity. The Hosmer-Lemeshow goodness-of-fit test was statistically significant (p=0.03), as commonly occurs at large sample sizes, while the C-statistic of 0.75 indicated acceptable discrimination. Adjusted odds ratios for all covariates are shown in Table 2 and Fig 2.

**Fig 2.**
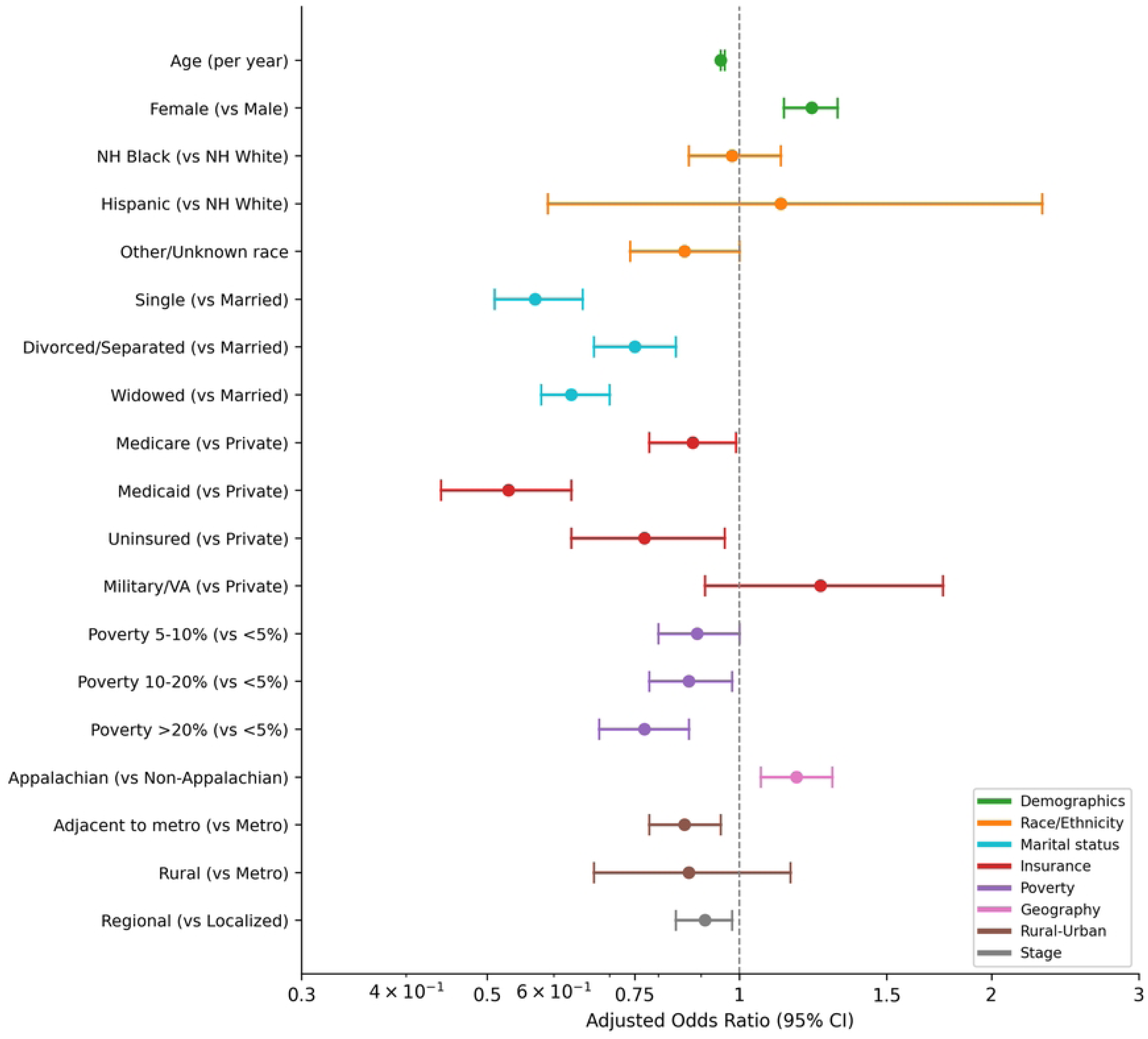
Forest Plot - Adjusted Odds Ratios for Guideline-Concordant Therapy Receipt (N=25,452)

### Interaction Effects

Three of four pre-specified interactions were significant at the α=0.10 level. The poverty × insurance interaction (p=0.009) suggested that poverty effects were amplified among underinsured patients. The Appalachian × poverty interaction (p=0.040) indicated that the poverty gradient in therapy receipt differed between Appalachian and non-Appalachian regions. The Appalachian × year interaction (p<0.001) indicated that the relationship between Appalachian status and therapy receipt changed over the study period (see Temporal Trends). The poverty × race interaction was not significant (p=0.536).

### Surgery versus Radiation (Secondary Model)

Among 22,375 patients receiving local therapy, predictors of receiving surgery versus radiation were similar but stronger. Poverty >20% was associated with 33% lower odds of surgery (OR 0.67, 95% CI 0.59-0.75), Medicaid with 43% lower odds (OR 0.57, 95% CI 0.48-0.68), and Military/VA insurance with 67% lower odds (OR 0.33, 95% CI 0.25-0.44). The C-statistic was 0.81, reflecting the strong association between age and treatment modality selection.

### Survival Analysis (Table 3)

Median overall survival (60-month cap) varied markedly by treatment: surgery 41.5 months, radiation 18.9 months, systemic only 12.3 months, and no cancer-directed therapy 7.7 months (log-rank p<0.001). By Appalachian status, median survival was similar: 25.0 months (Appalachian) versus 25.6 months (non-Appalachian; log-rank p=0.002). Poverty was associated with a stepwise decrease in survival (<5%: 28.0 months; >20%: 23.5 months; p<0.001).

**Table 3.** Adjusted Hazard Ratios for All-Cause Mortality (N=25,452)

| Group | Variable | HR | 95% CI | P |
| --- | --- | --- | --- | --- |
| Treatment | Radiation (ref: Surgery) | 2.12 | 2.02-2.22 | <0.001 |
| Treatment | Systemic only | 3.43 | 3.14-3.74 | <0.001 |
| Treatment | No cancer-directed therapy | 4.33 | 4.10-4.56 | <0.001 |
| Demographics | Age (per year) | 1.02 | 1.01-1.02 | <0.001 |
| Demographics | Female (ref: Male) | 0.78 | 0.75-0.81 | <0.001 |
| Race/Ethnicity | NH Black (ref: NH White) | 0.87 | 0.81-0.93 | <0.001 |
| Race/Ethnicity | Hispanic | 1.08 | 0.80-1.46 | 0.604 |
| Race/Ethnicity | Other/Unknown | 1.25 | 1.16-1.35 | <0.001 |
| Marital Status | Single (ref: Married) | 1.09 | 1.03-1.17 | 0.007 |
| Marital Status | Divorced/Separated | 1.08 | 1.02-1.14 | 0.007 |
| Marital Status | Widowed | 1.11 | 1.06-1.17 | <0.001 |
| Insurance | Medicare (ref: Private) | 1.14 | 1.07-1.20 | <0.001 |
| Insurance | Medicaid | 1.25 | 1.14-1.37 | <0.001 |
| Insurance | Uninsured/Self-pay | 1.07 | 0.97-1.19 | 0.178 |
| Poverty Index | 5-10% (ref: <5%) | 1.04 | 0.98-1.10 | 0.170 |
| Poverty Index | 10-20% | 1.06 | 1.00-1.12 | 0.033 |
| Poverty Index | >20% | 1.13 | 1.07-1.20 | <0.001 |
| Geography | Appalachian (ref: Non-Appalachian) | 1.07 | 1.02-1.12 | 0.004 |
| Rural-Urban | Adjacent to metro (ref: Metro) | 0.99 | 0.94-1.04 | 0.607 |
| Rural-Urban | Rural | 1.10 | 0.97-1.26 | 0.142 |
| Stage | Localized (ref: Regional) | 0.69 | 0.67-0.72 | <0.001 |
| Tumor | Tumor size (per mm) | 1.00 | 1.00-1.00 | <0.001 |

Concordance index = 0.711. Survival capped at 60 months. HR = hazard ratio; CI = confidence interval.

In multivariable Cox regression (concordance index 0.711, N=25,452), the strongest predictor of mortality was no cancer-directed therapy (HR 4.33, 95% CI 4.10-4.56 vs surgery). After adjustment for treatment and all covariates, poverty >20% remained significantly associated with worse survival (HR 1.13, 95% CI 1.07-1.20), as did Medicaid insurance (HR 1.25, 95% CI 1.14-1.37). Appalachian residence was associated with modestly worse all-cause survival (HR 1.07, 95% CI 1.02-1.12; p=0.004), an association that attenuated to the null in the cancer-specific competing-risks analysis (Fine-Gray subdistribution hazard ratio 1.01, 95% CI 0.96-1.06).

Schoenfeld residual testing indicated a violation of the global proportional hazards assumption (p<0.001), driven principally by treatment category, age, stage, tumor grade, smoking, and insurance; the socioeconomic exposures of interest (county poverty, Appalachian residence, race/ethnicity, and rurality) did not violate the assumption (all p>0.10). Reported hazard ratios are therefore interpreted as effects averaged over the 60-month follow-up. Consistent with residual immortal-time and confounding-by-indication effects, the hazard ratio for no cancer-directed therapy attenuated from 4.33 to 2.96 (95% CI 2.77-3.17) in a 6-month landmark analysis (N=17,679, excluding 9,077 patients who died within 6 months of diagnosis), while the socioeconomic associations remained essentially unchanged. Adjusted hazard ratios for all covariates are shown in Table 3 and Fig 3.

**Fig 3.**
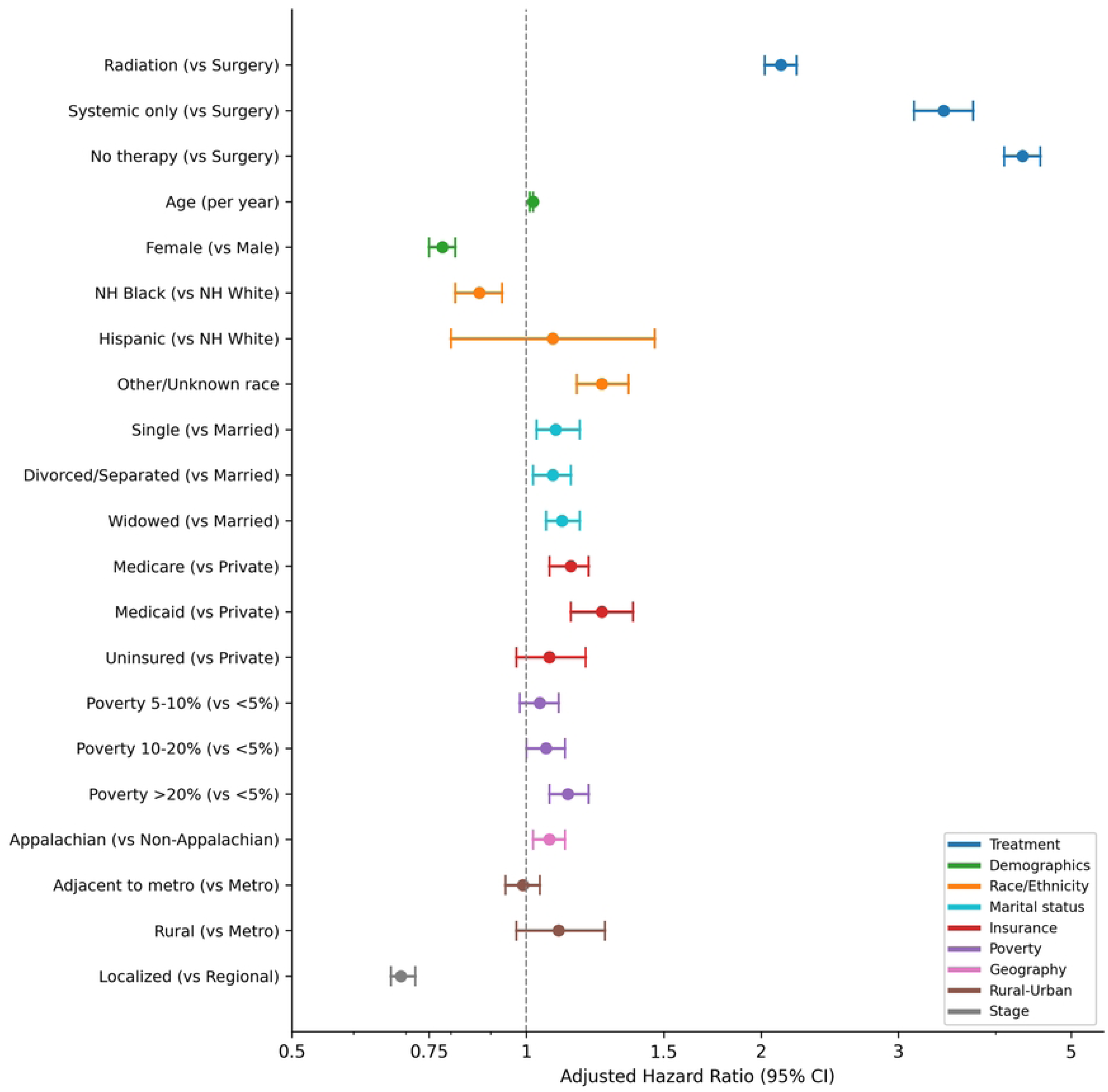
Forest Plot - Adjusted Hazard Ratios for All-Cause Mortality (N=25,452)

Patient and tumor characteristics differed across the four treatment categories (Table 4): patients who received no cancer-directed therapy were older (median 76 vs 69 years for surgical patients) and more often widowed (34.4% vs 17.8%), consistent with the survival differences described above.

**Table 4.** Patient and Tumor Characteristics by Treatment Category (N=26,756)

| Variable | Level | Surgery<br>(N=15,941) | Radiation<br>(N=6,434) | Systemic<br>(N=753) | No Therapy<br>(N=3,628) | p |
| --- | --- | --- | --- | --- | --- | --- |
| Age, years | Median (IQR) | 69 (61-75) | 74 (66-80) | 72 (64-78) | 76 (68-82) | <0.001 |
| Sex | Male | 7,765<br>(48.7%) | 3,420<br>(53.2%) | 410 (54.4%) | 1,869<br>(51.5%) | <0.001 |
|  | Female | 8,176 (51.3%) | 3,013<br>(46.8%) | 343 (45.6%) | 1,759<br>(48.5%) |  |
| Race/Ethnicity | NH White | 13,399<br>(84.1%) | 5,414<br>(84.1%) | 635 (84.3%) | 2,974<br>(82.0%) | <0.001 |
|  | NH Black | 1,402 (8.8%) | 690 (10.7%) | 69 (9.2%) | 410 (11.3%) |  |
|  | Hispanic | 57 (0.4%) | 23 (0.4%) | 2 (0.3%) | 9 (0.2%) |  |
|  | Other/Unknown | 1,083 (6.8%) | 307 (4.8%) | 47 (6.2%) | 235 (6.5%) |  |
| Marital Status | Married/Partnered | 9,002<br>(57.9%) | 3,068<br>(48.9%) | 391 (53.9%) | 1,370<br>(39.5%) | <0.001 |
|  | Single | 1,623 (10.4%) | 651 (10.4%) | 71 (9.8%) | 429 (12.4%) |  |
|  | Divorced/Separated | 2,154 (13.9%) | 923 (14.7%) | 90 (12.4%) | 477 (13.7%) |  |
|  | Widowed | 2,766 (17.8%) | 1,632<br>(26.0%) | 174 (24.0%) | 1,194<br>(34.4%) |  |
| Insurance | Private | 3,611<br>(23.1%) | 742 (11.7%) | 99 (13.5%) | 337 (9.4%) | <0.001 |
|  | Medicare | 9,735 (62.3%) | 4,704<br>(74.3%) | 520 (70.7%) | 2,714<br>(76.1%) |  |
|  | Medicaid | 930 (6.0%) | 357 (5.6%) | 44 (6.0%) | 240 (6.7%) |  |
|  | Uninsured/Self-pay | 872 (5.6%) | 188 (3.0%) | 47 (6.4%) | 104 (2.9%) |  |
| Military/VA | 154 (1.0%) | 181 (2.9%) | 5 (0.7%) | 60 (1.7%) |  |  |
| Unknown | 324 (2.1%) | 159 (2.5%) | 20 (2.7%) | 112 (3.1%) |  |  |
| Smoking | Never | 641 (4.0%) | 117 (1.8%) | 42 (5.6%) | 165 (4.5%) | <0.001 |
| Former | 3,112 (19.5%) | 1,170 (18.2%) | 140 (18.6%) | 615 (17.0%) |  |  |
| Current | 3,118 (19.6%) | 1,026 (15.9%) | 150 (19.9%) | 605 (16.7%) |  |  |
| Unknown | 9,070 (56.9%) | 4,121 (64.1%) | 421 (55.9%) | 2,243 (61.8%) |  |  |
| Poverty Index | <5% | 2,752 (17.3%) | 968 (15.1%) | 132 (17.6%) | 509 (14.1%) | <0.001 |
| 5-10% | 4,258 (26.8%) | 1,533 (23.9%) | 199 (26.5%) | 863 (23.9%) |  |  |
| 10-20% | 5,075 (31.9%) | 2,090 (32.6%) | 215 (28.6%) | 1,175 (32.5%) |  |  |
| >20% | 3,813 (24.0%) | 1,822 (28.4%) | 205 (27.3%) | 1,066 (29.5%) |  |  |
| Rural-Urban | Metro | 12,973 (81.4%) | 5,228 (81.3%) | 611 (81.1%) | 2,891 (79.7%) | 0.416 |
| Adjacent to metro | 2,676 (16.8%) | 1,092 (17.0%) | 130 (17.3%) | 666 (18.4%) |  |  |
| Rural | 292 (1.8%) | 114 (1.8%) | 12 (1.6%) | 71 (2.0%) |  |  |
| Stage | Localized | 11,747 (73.7%) | 4,395 (68.3%) | 448 (59.5%) | 2,650 (73.0%) | <0.001 |
| Regional (direct ext.) | 4,194 (26.3%) | 2,039 (31.7%) | 305 (40.5%) | 978 (27.0%) |  |  |
| Tumor Size, mm | Median (IQR) | 23 (15-35) | 29 (19-44) | 35 (23-54) | 30 (20-46) | <0.001 |
| Grade | Well differentiated | 2,535 (15.9%) | 375 (5.8%) | 66 (8.8%) | 256 (7.1%) | <0.001 |
| Moderately differentiated | 6,470 (40.6%) | 1,059 (16.5%) | 123 (16.3%) | 519 (14.3%) |  |  |
| Poorly differentiated | 4,900 (30.7%) | 1,506 (23.4%) | 171 (22.7%) | 691 (19.0%) |  |  |
| Undifferentiated | 234 (1.5%) | 83 (1.3%) | 12 (1.6%) | 35 (1.0%) |  |  |
| Unknown | 1,802 (11.3%) | 3,411 (53.0%) | 381 (50.6%) | 2,127 (58.6%) |  |  |
| Laterality | Right | 9,476 (59.5%) | 3,520 (55.1%) | 395 (53.2%) | 1,972 (54.7%) | <0.001 |
| Left | 6,439 (40.5%) | 2,871 (44.9%) | 347 (46.8%) | 1,631 (45.3%) |  |  |
| Diagnosis Year | 2004-2006 | 4,047 (25.4%) | 1,266 (19.7%) | 216 (28.7%) | 873 (24.1%) | <0.001 |
| 2007-2009 | 3,958 (24.8%) | 1,323 (20.6%) | 184 (24.4%) | 879 (24.2%) |  |  |
| 2010-2012 | 3,836 (24.1%) | 1,706<br>(26.5%) | 197 (26.2%) | 927 (25.6%) |  |  |
| 2013-2015 | 4,100 (25.7%) | 2,139<br>(33.2%) | 156 (20.7%) | 949 (26.2%) |  |  |

*Radiation = radiation therapy without surgery; Systemic = systemic therapy alone. Values are n (%) or median (IQR); p-values from chi-squared (categorical) or Wilcoxon rank-sum (continuous) tests. Percentages are computed over non-missing observations within each treatment column.*

### Temporal Trends

Guideline-concordant therapy rates converged between Appalachian and non-Appalachian regions over the study period. The adjusted Appalachian OR shifted from 1.38 (95% CI 1.16-1.64) in 2004-2007 to 1.27 (95% CI 1.08-1.51) in 2008-2011, and was no longer significant by 2012-2015 (OR 0.92, 95% CI 0.78-1.08; year × Appalachian interaction p<0.001). Surgery rates declined in both regions over this period, with Appalachian counties declining from 64.3% to 55.5% and non-Appalachian from 62.3% to 55.8%, consistent with the national trend toward increased use of stereotactic radiation for early-stage disease.

### Sensitivity Analyses

E-values for the primary findings were: Medicaid OR 0.53 (E-value 3.20 for the point estimate and 2.55 for the confidence limit nearest the null) and poverty >20% OR 0.77 (E-value 1.91; CI limit 1.55). Because the odds ratio overstates the risk ratio at this high outcome prevalence, these odds-ratio E-values are upper bounds on robustness, and the corresponding risk-ratio effects (S1 Table) would yield smaller values. The Medicaid effect would nonetheless require an unmeasured confounder associated with both insurance and treatment by a factor of 2.55 to 3.20, above and beyond measured covariates, to explain it away, a magnitude exceeding that of most measured confounders in cancer disparities research.

The 2010-2015 restricted analysis (N=14,010) produced highly consistent results: poverty >20% OR 0.76 (95% CI 0.63-0.91), Medicaid OR 0.59 (95% CI 0.46-0.76). Race-stratified models revealed that the poverty gradient was significant only among non-Hispanic White patients (poverty >20% OR 0.75, p<0.001) and not among non-Hispanic Black patients (OR 1.02, p=0.93). Medicaid was significantly associated with reduced therapy in both groups (White OR 0.53; Black OR 0.48).

A sensitivity analysis restricted to potentially operable candidates, excluding the 1,590 non-surgical patients with a documented comorbidity contraindication or death before treatment (operable-candidate cohort N=25,166; multivariable model N=23,946 after complete-case exclusions), yielded materially unchanged socioeconomic associations (Medicaid OR 0.52, 95% CI 0.43-0.62; county poverty >20% OR 0.78, 95% CI 0.68-0.89; Appalachian OR 1.15, 95% CI 1.04-1.28), confirming that the observed disparities are not attributable to documented medical inoperability (S2 Table).

IPTW-weighted Cox analysis showed a strong association between guideline-concordant therapy and survival (HR 0.35, 95% CI 0.33-0.37; propensity score range 0.20 to 0.99 with adequate overlap between groups, stabilized weights trimmed at the 1st and 99th percentiles to 0.31-2.54). Because performance status and comorbidity are not captured in the registry, this estimate is interpreted as an adjusted association rather than a causal effect. Poverty effects on survival remained significant after propensity weighting for treatment receipt.

In a strict complete-case analysis (N=7,208, 27% of the cohort, restricted to patients with documented smoking and grade), the direction of all key associations was preserved, although magnitudes shifted and precision fell: the Medicaid association strengthened (OR 0.32, 95% CI 0.19-0.55) while the county-poverty gradient attenuated and was no longer statistically significant (poverty >20% OR 0.83, 95% CI 0.61-1.13). A multiple-imputation analysis (chained equations, m=10) imputing smoking and tumor grade yielded results consistent with the primary model (Medicaid OR 0.49, 95% CI 0.42-0.59; poverty >20% OR 0.77, 95% CI 0.69-0.87; Appalachian OR 1.17, 95% CI 1.06-1.28; S2 Table), indicating that the principal findings are robust to the handling of missing data, with the poverty gradient most sensitive to the loss of power in the smaller complete-case subset.

A modified Poisson regression with robust standard errors[22] yielded adjusted prevalence ratios consistent in direction and statistical significance with the primary logistic odds ratios; the magnitude of associations was attenuated on the risk-ratio scale, as expected at the observed high (approximately 84%) outcome prevalence. Medicaid insurance (RR 0.94, 95% CI 0.91-0.96), county-level poverty >20% (RR 0.96, 95% CI 0.95-0.98), and Appalachian residence (RR 1.02, 95% CI 1.01-1.04) remained statistically significant; adjusted prevalence ratios for all covariates are reported in S1 Table.

On the absolute scale, model-standardized predicted probabilities from the primary model indicated that the probability of receiving guideline-concordant therapy was 85.5% under universal private insurance versus 77.1% under universal Medicaid (adjusted difference 8.3 percentage points), and 85.5% versus 82.5% comparing county poverty <5% with >20% (difference 3.0 percentage points; S2 Table). In competing-risks analyses of cancer-specific mortality (treating non-cancer death as a competing event), the socioeconomic associations persisted (Medicaid Fine-Gray subdistribution hazard ratio 1.22, 95% CI 1.11-1.34; county poverty >20% 1.10, 95% CI 1.03-1.18), whereas Appalachian residence remained null (1.01, 95% CI 0.96-1.06; S3 Table).

## DISCUSSION

This statewide, population-based analysis of 26,756 patients with early-stage NSCLC demonstrates that socioeconomic factors, particularly insurance status and poverty, rather than Appalachian geography itself, are the patient factors most strongly associated with disparities in receipt of guideline-concordant treatment in Ohio. Several findings merit emphasis.

### Insurance as the Dominant Barrier

Medicaid insurance emerged as the strongest modifiable predictor of failure to receive guideline-concordant local therapy, conferring 47% lower adjusted odds compared with private insurance (OR 0.53, 95% CI 0.44-0.63). Because the odds ratio overstates the risk ratio when the outcome is common, the effect is more conservatively expressed on the absolute and risk-ratio scales: model-standardized predictions from the primary model indicate roughly 8 fewer guideline-concordant treatments per 100 patients (85.5% under universal private insurance versus 77.1% under universal Medicaid), and the modified-Poisson adjusted risk ratio (0.94; S1 Table) likewise shows attenuation relative to the odds ratio. This association was consistent across time periods, racial strata, multiple imputation, and an operable-candidates sensitivity analysis, and was supported by an E-value of 2.55 to 3.20, indicating meaningful robustness to unmeasured confounding. The magnitude of the insurance association exceeded that of poverty, race, or geography, positioning insurance status as the most actionable target for intervention.

This finding closely mirrors results from Stokes et al., who reported an identical Medicaid odds ratio (0.53) in a national NCDB analysis of 240,361 patients with early-stage NSCLC.[6] Our study extends these observations by using a comprehensive population-based registry that captures all diagnosed patients, including those who never access Commission on Cancer-accredited facilities, thereby enhancing generalizability. The importance of insurance coverage is further supported by evidence from Medicaid expansion studies: Zhao et al. demonstrated that Medicaid expansion was associated with a 2.10 percentage-point increase in timely guideline-concordant therapy and a 1.79 percentage-point improvement in 5-year survival among patients with NSCLC.[24] However, Gawdi et al. showed that residents of the poorest counties, particularly in rural and small-metropolitan areas, did not experience proportional survival gains following expansion,[29] suggesting that insurance expansion alone may be insufficient to overcome entrenched structural barriers in high-poverty communities.

### Poverty Effects and Racial Heterogeneity

County-level poverty was independently associated with reduced receipt of guideline-concordant therapy, with patients residing in counties with >20% poverty experiencing a 23% reduction in treatment odds and a clear dose-response gradient across poverty categories. These findings align with national evidence documenting substantial geographic variation in surgical treatment, with low-receipt counties characterized by concentrated poverty, high uninsured rates, and nonmetropolitan status.[30] Consistent with prior work by Ebner et al., our results also highlight the cumulative disadvantage posed by socioeconomic risk clustering.[7]

Notably, the observed poverty gradient was confined to non-Hispanic White patients. In race-stratified models, non-Hispanic White patients showed a significant gradient (OR 0.75 for >20% poverty, p<0.001), whereas non-Hispanic Black patients exhibited no association between county-level poverty and therapy receipt (OR 1.02, p=0.93). This pattern may reflect differing mechanisms of access: for Black patients, systemic barriers such as residential segregation and institutional access differences may operate relatively independently of income gradients,[23] whereas for White patients, poverty may represent a more differentiating determinant of care access. However, these findings should be interpreted cautiously: non-Hispanic Black patients comprised only 9.6% of the cohort, and the formal poverty × race interaction was not statistically significant (p=0.536). The divergent patterns are consistent with prior literature but require confirmation in larger cohorts with greater racial diversity.

### Geography, Socioeconomic Composition, and Access

A central finding of this study is that Appalachian residence was not independently associated with lower receipt of guideline-concordant therapy; after adjustment for insurance, poverty, and other socioeconomic factors, Appalachian residents were if anything modestly more likely to receive it (OR 1.17; adjusted risk ratio 1.02). This contrasts with findings from Tennessee, where Mamudu et al. reported a 35% reduction in surgical treatment odds for residents of Appalachian counties.[14] Such discrepancies likely reflect state-specific differences in healthcare infrastructure and referral networks; notably, the Appalachian counties contributing to this cohort are served by a Commission on Cancer-accredited regional center, which may account for the absence of a geographic deficit and cautions against generalizing this directional finding beyond Ohio.

Prior work by Vanderpool et al. has demonstrated substantial heterogeneity in cancer-related beliefs and behaviors across Appalachian states, reinforcing that “Appalachian” identity does not represent a uniform exposure.[31] Our results indicate that in Ohio, lower therapy receipt is concentrated among underinsured and high-poverty populations rather than determined by Appalachian geography: insurance and poverty were independently associated with treatment receipt, whereas Appalachian residence was not a barrier. This distinction has direct policy relevance, as interventions targeting Appalachian designation alone may be less effective than those addressing the underlying socioeconomic disadvantage that is unevenly distributed both within and outside Appalachian counties.

Temporal analyses further underscore this point. Rates of guideline-concordant therapy converged between Appalachian and non-Appalachian regions over time, with no meaningful differences by 2012-2015. This convergence coincided with declining surgical resection rates in both regions, paralleling national adoption of SBRT for early-stage disease. Ganesh et al. demonstrated that rising SBRT utilization (from 1% to 22% nationally) reduced no-treatment rates from 22% to 10.5% and narrowed racial treatment gaps.[25] Ray et al. showed that receipt of any guideline-concordant local therapy, regardless of modality, eliminated rurality-associated survival disparities,[32] a finding supported by other analyses suggesting that ensuring treatment access rather than dictating modality is the critical intervention.[33] The significant year × Appalachian interaction (p<0.001) highlights the importance of longitudinal analyses, as cross-sectional assessments may overstate or misattribute geographic disparities.

### Reframing the “No Treatment” Narrative

An important contribution of this study is the comprehensive classification of cancer-directed treatment. Analyses that count only surgical resection can suggest that a large minority of patients with early-stage disease receive no curative treatment; by classifying both radiation (24.0%) and systemic therapy (2.8%) as active cancer-directed treatment, we found a no-therapy rate of 13.6% in this cohort. This is lower than the approximately 22% no-treatment rate reported nationally before the widespread diffusion of stereotactic body radiation therapy, which subsequently declined to about 10.5%.[25] Thus, the central issue is not a universal absence of care, but disparities in the type and adequacy of care received.

Analysis of the ReasonNoSurg variable provides further insight. Among patients who did not undergo resection, surgery was not recommended in 73.6% of cases (likely reflecting medical inoperability or physician judgment), contraindicated due to comorbidity in 14.3%, and refused by 4.8%. Refusal rates did not differ meaningfully by Appalachian status (5.1% vs 4.7%), and the difference by race (4.6% non-Hispanic White vs 5.7% non-Hispanic Black) was directionally consistent with but substantially smaller in magnitude than national findings by Coaston et al., who reported that Black race was associated with doubled refusal odds (aOR 2.06).[17] The overall low refusal rates in this Ohio cohort suggest that patient preference is unlikely to be a primary driver of geographic disparities in surgical treatment. Rapp et al. demonstrated that surgery refusal contributes measurably to racial survival disparities nationally;[18] the comparatively modest racial gap in refusal observed here may reflect both the predominantly non-Hispanic White composition of Ohio’s Appalachian counties and the relatively small Black subsample (9.6% of the cohort).

### Limitations

Several limitations warrant acknowledgment. First, OCISS does not include performance status, pulmonary function testing, or a validated comorbidity index (Charlson or Elixhauser). These unmeasured confounders could partially explain treatment disparities, as sicker patients may be less likely to receive surgery. However, E-values indicate that a confounder would need a 3.2-fold association with both insurance status and treatment to fully explain the Medicaid effect, a magnitude exceeding most measured confounders. The ReasonNoSurg variable itself, although NAACCR-standardized, depends on physician documentation in the medical record and may incompletely distinguish true medical contraindication from physician judgment or patient-level barriers; accordingly, we restricted a sensitivity analysis to potentially operable candidates by excluding patients with a documented comorbidity contraindication or death before treatment, and the socioeconomic associations persisted (Results), indicating they are not attributable to patients with non-modifiable surgical barriers.

Second, smoking history was missing for 59.3% of patients and tumor grade for 28.9%. We addressed this through “Unknown” category coding rather than exclusion, which was validated by a complete-case sensitivity analysis showing consistent results. The high proportion of missing smoking data reflects evolving registry collection practices during the study period and is consistent with other state registry analyses.[34]

Third, county-level poverty is an ecologic measure that may not accurately reflect individual socioeconomic status. However, area-level deprivation indices have been validated as independent predictors of cancer outcomes.[35]

Fourth, the study period (2004-2015) precedes the widespread adoption of immunotherapy and lung cancer screening. Contemporary treatment patterns may differ, and these findings should be confirmed in more recent cohorts. Liu et al. documented persistent disparities even after SBRT became widely available,[36] suggesting that structural barriers may continue to operate regardless of therapeutic advances.

Fifth, OCISS does not capture hospital-level characteristics, including Commission on Cancer accreditation status, surgical volume, or facility type. Receipt of guideline-concordant therapy is known to vary by these factors, and unmeasured differences in healthcare delivery infrastructure across Ohio counties may contribute to residual variation in treatment patterns not fully explained by patient-level socioeconomic measures.

### Clinical and Policy Implications

These findings suggest several actionable strategies. First, although Medicaid insurance might be interpreted as a marker of socioeconomic disadvantage, it is itself a directly modifiable policy variable: state Medicaid expansion under the Affordable Care Act has been associated with both increased timely receipt of guideline-concordant lung cancer care and improved survival.[24] Ohio expanded Medicaid in 2014, late in our study window, preventing direct assessment of its impact on the cohort; the Gawdi et al. analysis found that residents of high-poverty counties realized smaller gains from expansion,[29] underscoring the need for parallel investment in cancer-care navigation, transportation, and care-coordination infrastructure to translate insurance coverage into actual receipt of therapy. Second, outreach and care-navigation efforts should prioritize socioeconomic risk rather than Appalachian designation alone, as poverty and insurance status, not geography, drive the observed gaps. Third, the observed decline in surgical resection across all regions warrants continued monitoring to ensure that increasing reliance on radiation therapy reflects appropriate clinical selection rather than diminished surgical access.

## Conclusions

In this population-based analysis of early-stage NSCLC in Ohio, insurance status (particularly Medicaid) and county-level poverty emerged as the strongest modifiable factors associated with receipt of guideline-concordant local therapy, while Appalachian residence was not a barrier to treatment; its modest association with all-cause survival (HR 1.07) attenuated to the null in a competing-risks analysis of cancer-specific mortality after accounting for socioeconomic context. These findings argue against framing lung cancer treatment disparities in purely geographic terms and instead support policy approaches that directly address insurance-related and poverty-related barriers to care in high-risk communities.

## Data Availability

The individual-level datasets analyzed in this study are restricted under Ohio Revised Code Section 3701.262 and cannot be publicly shared by the authors. Researchers may apply for access through the Ohio Cancer Incidence Surveillance System (OCISS) Data Use Application process, administered by the Ohio Department of Health Bureau of Vital Statistics application materials and the OCISS Data Request Committee contact information are available at https://odh.ohio.gov/know-our-programs/ohio-cancer-incidence-surveillance-system/data-requests. The authors received the analytic dataset under a signed Data Use Agreement and are not permitted to redistribute individual-level records. The analytic code (R scripts) used to produce all tables and figures is provided as Supporting Information (S1 Code).

## ACKNOWLEDGMENTS

The authors gratefully acknowledge the Ohio Department of Health Bureau of Vital Statistics, the Ohio Cancer Incidence Surveillance System data abstractors and staff, and the patients of Ohio whose records contributed to this analysis. This study includes data provided by the Ohio Department of Health, which should not be considered an endorsement of this study or its conclusions.

## DATA AVAILABILITY STATEMENT

The datasets generated and analyzed during the current study are restricted under Ohio Revised Code §3701.262 and are not publicly available. Researchers may apply for access through the Ohio Cancer Incidence Surveillance System Data Use Application process administered by the Ohio Department of Health Bureau of Vital Statistics; application materials and the OCISS Data Request Committee contact information are available at https://odh.ohio.gov/know-our-programs/ohio-cancer-incidence-surveillance-system/data-requests. The authors received the analytic dataset under a signed Data Use Agreement and are not permitted to redistribute individual-level records. The analytic code (R scripts) used to produce all tables and figures is provided as Supporting Information (S1 Code); aggregate, non-identifiable results underlying the figures are available from the corresponding author on request.

## SUPPORTING INFORMATION

**S1 Checklist.** STROBE Statement - checklist of items that should be included in reports of observational studies. (DOCX)

**S1 Code.** Analytic R scripts used to produce all tables, figures, and sensitivity analyses. (ZIP)

**S1 Table.** Adjusted prevalence ratios for receipt of guideline-concordant local therapy from a modified Poisson regression with robust standard errors (N=25,452). (DOCX)

**S2 Table.** Robustness of the socioeconomic associations: operable-candidate sensitivity analysis, multiple-imputation analysis, and adjusted predicted probabilities (g-computation). (DOCX)

**S3 Table.** Cancer-specific mortality: cause-specific and Fine-Gray competing-risks hazard ratios (60-month horizon). (DOCX)

## Notes

### Competing Interest Statement

The authors have declared no competing interest.

### Funding Statement

The author(s) received no specific funding for this work.

### Author Declarations

This study was approved under expedited review by the Ohio Department of Health Human Subjects Institutional Review Board (FWA00001963, IRB00002180 Protocol 2018-01). The analysis used de-identified secondary data from the Ohio Cancer Incidence Surveillance System informed consent was not obtained because the data were de-identified and analyzed anonymously.

